# A Reproducible Decomposed Large Language Model Pipeline for Structured Abstraction of Multimodal Neuroimaging Reports: Development and Independent Validation in 1,669 Emergency Department Encounters for Dizziness and Vertigo

**DOI:** 10.64898/2026.11.03.26367160

**Authors:** Jai Shahani, Abdul Rahman Ousta, Shaul Shaulov, Yasir El-Sherif

## Abstract

**Objective:** To develop and independently validate a decomposed, modality-specific large language model (LLM) pipeline that abstracts structured variables from the free-text neuroimaging reports of a multimodal stroke workup, and to validate the resulting structured dataset for reuse in downstream research.

**Materials and Methods:** A frozen pipeline (temperature 0) extracting 22 variables across CT head, CT angiography, CT perfusion, and MRI brain was applied to 1,669 ED encounters for dizziness or vertigo; 11 variables were pre-specified as primary (Tier 1). Reproducibility used three same-session runs and a nine-week re-run; three frontier models (Claude Sonnet 4.6, GPT-5.4, Gemini 3.5 Flash) measured cross-model agreement (Cohen’s/Fleiss’ κ). Accuracy was assessed in an independent 200-encounter cohort (150 random, 50 enriched for rare findings); 100 were double-read by two blinded reviewers with discordances adjudicated.

**Results:** Test–retest reproducibility was high (Tier 1 Fleiss’ κ=0.993; 99.8% exact) and stable nine weeks later (κ=0.992); inter-model agreement was κ=0.962. Against the adjudicated reference, pooled Tier 1 accuracy was 0.971 to 0.988 across models (Cohen’s κ 0.959 to 0.982), comparable to inter-reviewer agreement (κ=0.932). Accuracy was 0.980 to 0.983 in random and 0.955 to 0.977 in enriched encounters (agreement on model-flagged findings, not sensitivity).

**Discussion:** A decomposed LLM pipeline produced reproducible, model-portable, stable, and accurate multimodal abstraction. Because schema design and validation are up-front costs, such pipelines scale to larger research cohorts.

**Conclusion:** The quantified per-field error rates support reuse of this dataset with field-specific review, and such pipelines offer a practical path to large-scale, reproducible research datasets.

## Background and Significance

Turning free-text radiology reports into structured, analyzable variables is a persistent bottleneck in clinical research data curation. Manual abstraction is accurate but slow, requires domain expertise, and is reviewer-dependent, which in practice caps the size of cohorts that can be assembled and the number of questions that can be asked of them. This constraint is acute in the emergency evaluation of dizziness and vertigo, where distinguishing benign vestibular causes from posterior-circulation stroke frequently requires a multimodal workup: non-contrast CT head (CTH), CT angiography (CTA) of the head and neck, CT perfusion (CTP), and MRI brain. Each study generates a separate narrative report whose vessel-grading and posterior-fossa terminology must be interpreted and encoded.^1,2,3,4,5^ Dizziness and vertigo are among the most common emergency department presentations, at roughly 3 to 4% of visits, and a recognized source of missed posterior-circulation stroke, which makes accurate, scalable capture of this workup valuable.^6,7,8^

Classical natural-language-processing (NLP) systems automated parts of this task but remained narrow, single-task classifiers that required annotated training data and did not generalize across modalities.^9,10^ Large language models (LLMs) now extract structured variables from unstructured clinical text with accuracy that can match or exceed specialized tools, and prompt specificity, not model choice alone, largely governs quality, and open-source models achieve high agreement extracting from emergency neuroimaging reports specifically.^11,12,13,14,15^ LLM-based abstraction is accordingly being adopted for large-scale cohort and registry curation, with validation frameworks emerging to certify machine-extracted data.^16,17,18,19,20^ Three gaps recur, however: prompt and schema design are rarely standardized or reported; reproducibility, meaning whether a model returns the same answer on re-execution, and cross-developer agreement are seldom quantified, despite being prerequisites for trusting machine-extracted data; and most studies address a single modality, usually CT head alone. We are not aware of a prior pipeline that abstracts the full CTH, CTA, CTP, and MRI stroke workup at a scale approaching 1,669 encounters with reproducibility, cross-model, and independently adjudicated accuracy reporting in one system.

## Objective

We developed a decomposed, modality-specific LLM pipeline for structured abstraction of the multimodal stroke-workup neuroimaging cascade, and evaluated three properties required of a research-grade extraction tool: (1) reproducibility, both within a session and across a multi-week interval; (2) agreement across independently developed frontier models; and (3) field-level accuracy against an independent, two-reviewer, adjudicated human reference. We pre-specified eleven high-confidence variables as the primary (Tier 1) endpoints. The overarching aim was to validate the resulting dataset for reuse by quantifying, per field, the error a future study would inherit. Although instantiated for stroke imaging, the architecture (modality-specific decomposition with fixed schemas, deterministic quality control, and prevalence-informed sampling) is intended as a generalizable template for multimodal report abstraction.

## Materials and Methods

### Study design and cohort

This retrospective study at Staten Island University Hospital (Northwell Health) covered January 1, 2022– October 31, 2025 and was approved by the Northwell Health IRB (#26-0151; Category 5 expedited), with a waiver of consent and HIPAA authorization; all patient identifiers were removed before analysis. Reporting follows STARD 2015, adapted for automated report abstraction (checklist provided as a separate supplementary file). Adults (≥18 years) presenting to the ED with an initial complaint of dizziness or vertigo were eligible; all consecutive eligible encounters over the study period were extracted by the institutional quantitative-intelligence team, defining 1,669 encounters. Imaging reflects the institution’s usual stroke-workup pathway, not a study-imposed requirement. MRI completion was not an inclusion requirement; 719 encounters (43.1%) completed MRI and constitute the subset for MRI-dependent fields.

### Pipeline architecture (v14.0.3)

Each encounter passes through up to six single-turn LLM calls at temperature 0 (top_p 0.95 for GPT-5.4 and Gemini; Claude temperature 0 only; no fixed seed) via an institutional enterprise generative API: (1) CT head (acute-finding status, an indeterminate flag, acute-finding type, microvascular disease), which also receives the CTA head report so that non-contrast head findings described within a combined CTA report are captured; (2) CTA head/neck, grading seven vessels (bilateral vertebral, basilar, bilateral internal carotid, bilateral middle cerebral) on a fixed 0–4 stenosis scale (0 = none, 1 = <50%, 2 = 50–69%, 3 = 70–99%/near-occlusion, 4 = occlusion; 9 = not evaluable when no CTA text); (3) CT perfusion (technical quality, perfusion deficit), called only when a deterministic pattern match finds CT-perfusion terminology in the CTA reports (the same match sets CT-perfusion-performed status); (4) MRI core (acute infarct, chronic infarct, other abnormality and type, technical quality); and (5–6) acute- and chronic-infarct localization (territory, bilaterality), called only when the MRI core call returns a positive acute or chronic infarct. MRA inclusion is determined deterministically from MRI report text rather than by an LLM call. Calls 1–4 are independent of one another and each reads only source report text; the only LLM output passed to another LLM call is the infarct evidence phrase that the MRI core call supplies to the localization calls (together with the MRI report). No call uses tools, retrieval, memory, planning, or messaging between calls, and cross-modal information is combined only by the deterministic layers described next (Figure 1B). A per-case confidence score is computed deterministically from cross-modal concordance rules (for example, reconciling an indeterminate CT-head finding against a confirmatory MRI). A deterministic quality-control layer then scans report text with rule-based regular expressions (not a learned model) for unnegated abnormality language inconsistent with the structured output; 80 of 1,669 encounters (4.8%) in Claude run 1 (50 for GPT-5.4, 57 for Gemini) triggered at least one such flag; most applied a value-changing correction, and the remainder were advisory flags that recorded a possible inconsistency (for example, unnegated abnormality wording, or a vessel grade lacking a supporting phrase) without altering the value. A JSON-repair and exponential-backoff layer ensured near-complete processing. All prompts, rules, and thresholds were frozen in v14.0.3 (July 22, 2026) before any production run and before validation sampling. Models were called as claude-sonnet-4.6, gpt-5.4, and gemini-3.5-flash; no dated snapshot identifier was available, so run dates are reported (production runs August 1–5, 2026; GPT-5.4 re-run October 8, 2026). Prompts and code are provided (Data availability).

**Figure 1.**
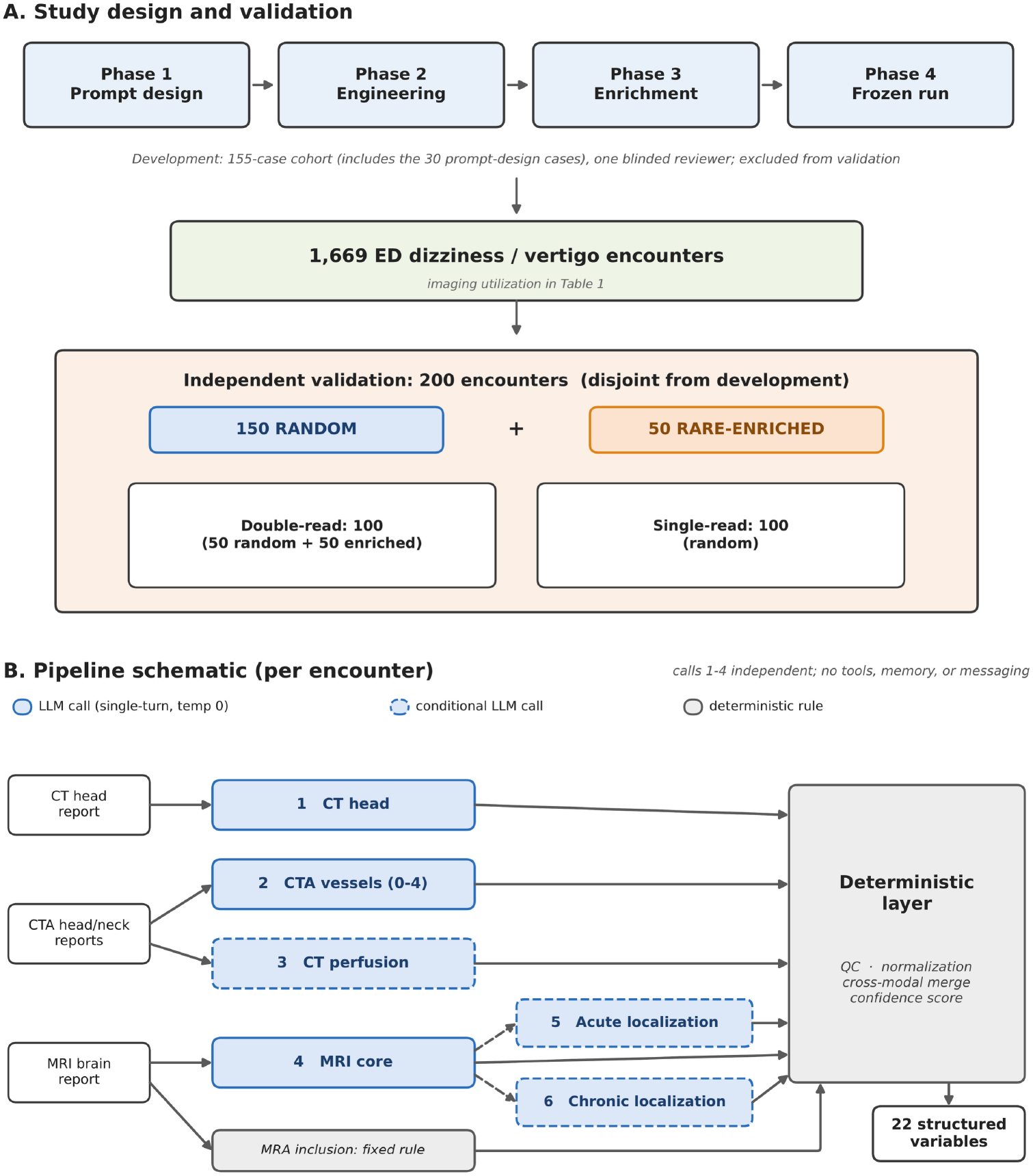
(A) Study design. The pipeline was developed over four phases (prompt design, prompt engineering across a four-model panel, prevalence-informed enrichment for rare findings, and the frozen production run), producing a 155-case development cohort (112 random, including the 30 initial prompt-design cases, plus 43 curated) that was coded by a single blinded development reviewer and excluded from validation. The frozen pipeline (v14.0.3) was applied to all 1,669 encounters (imaging utilization in Table 1). Accuracy was established against 200 encounters disjoint from development: 150 sampled at random and 50 prevalence-enriched from frozen model output to over-sample rare findings, interleaved with their origin concealed from reviewers. Of the 200, 100 (50 random and 50 enriched) were double-read by two blinded neurology residents (inter-rater κ=0.93) with discordances resolved by a blinded adjudicator to form the reference standard; the remaining 100 random encounters were single-read (50 per reviewer, that reviewer’s coding serving as the reference). In total, 355 encounters underwent expert manual review. (B) Pipeline schematic (v14.0.3). Up to six single-turn LLM calls run at temperature 0: (1) CT head, which also reads the CTA head text; (2) CTA vessel grading of seven vessels on a 0–4 scale; (3) CT perfusion, gated by a regular-expression match; (4) MRI core (acute infarct, chronic infarct, other abnormality, quality); and (5–6) acute- and chronic-infarct localization, each called only when the MRI core call returns the corresponding infarct. A deterministic layer then applies rule-based quality-control checks, grade normalization, cross-modal merging (for example, an MRI resolving an indeterminate CT-head finding), and a confidence score, and MR angiography inclusion is set by a fixed rule on the MRI report; the pipeline outputs 22 structured variables. Calls 1–4 are independent; the only LLM-to-LLM hand-off is the MRI evidence phrase passed to the localization calls. No call uses tools, memory, retrieval, or messaging.

### Variable tiers

The 22 variables were pre-specified into eleven primary (Tier 1) and eleven secondary (Tier 2) fields before validation (finalized before validation, not preregistered). Tier 1 comprised the CTH acute finding and microvascular disease; the seven CTA vessel grades; whether CTP was performed; and acute infarct on MRI, which are the findings most consequential to the dizziness/stroke decision and most cleanly defined by the categorical schema. Each vessel was graded on a 0–4 stenosis scale; a value of 9 is assigned only when no CTA report text is available (11 encounters) and was treated as not evaluable (excluded from numerators and denominators); within a reported CTA, a vessel that is not described is graded 0. Agreement and accuracy were computed on the full 0–4 categorical grade; finding prevalence is reported as the proportion of assessable vessels with any stenosis (grade ≥1).

**Table 1.** Imaging utilization across the 1,669-encounter cohort. CT head, CTA head, CTA neck, and MRI brain performance are drawn from structured study-performed fields. CT perfusion and MR angiography have no dedicated study-performed fields in this dataset and were ascertained by the pipeline from report text. CT perfusion is listed among the studies ordered and reported together with CTA head and CTA neck, so it was ascertainable for the full cohort (denominator 1,669). MR angiography could be identified only from MRI brain reports and is therefore expressed among the 719 MRI encounters. CT-perfusion-performed and MR-angiography-inclusion status were set by deterministic text rules (CT perfusion is listed with the CTA reports; MR angiography appears only in MRI reports); the gated CT-perfusion LLM call extracted the remaining perfusion fields. As an implementation check, the rule-based CT-perfusion flag was consistent with the report text in 1,668/1,669 encounters; human-reference accuracy for these fields is reported in the Supplement.

| **Imaging study** | **n performed** | **%** | **Source** |
|---|---|---|---|
| CT head (CTH) | 1,650 | 98.9% | Structured EHR field |
| CTA head | 1,666 | 99.8% | Structured EHR field |
| CTA neck | 1,661 | 99.5% | Structured EHR field |
| MRI brain | 719 | 43.1% | Structured EHR field |
| CT perfusion | 449 | 26.9% | CTA report text |
| MR angiography | 27 | 3.8%* | MRI report text |

*Percentages are of the 1,669-encounter cohort except where marked. \*MR angiography could be ascertained only from MRI brain reports: 27/719 MRI encounters (3.8%); 1.6% of the full cohort.*

### Iterative development and the 155-case development cohort

The pipeline was refined over four phases. Phase 1 began with 30 cases, sampled at random from the MRI cohort, for prompt design; these 30 are the first cases of the 155-case development cohort described below and, like all development cases, were excluded from the validation sample. Phase 2 performed prompt engineering across a four-model panel (GPT-5, GPT-5-mini, Gemini 2.5 Pro, and Gemini 2.5 Flash), driven by multi-model concordance rather than human review. A first batch of 75 cases was then reviewed once by the development reviewer (a general neurologist and director of a Joint Commission-certified primary stroke program, separate from the three validation reviewers), confirming high accuracy on common findings but sparse coverage of rare endpoints. In Phase 3, multi-model consensus over the 719 MRI encounters was treated as a proxy for prevalence to size a targeted, prevalence-informed enrichment (target ≥14 human-confirmed positives per endpoint, for an approximately ±15-point 95% Wilson-score interval), producing a 155-case development cohort (112 random, which include the 30 initial prompt-design cases, plus 43 curated) drawn from the MRI cohort. This exhausted the model-flagged hemorrhages in the MRI cohort (all 18 hemorrhage encounters flagged by Claude among the 719 were in the 155), so the subsequent enrichment of the validation sample (below) drew on all 1,669 encounters. This multi-model consensus was only a sampling signal; no accuracy estimate used it, all being computed against the human-adjudicated reference. In Phase 4 the frozen architecture was run across the full cohort with three frontier models. The development reviewer coded the 155 development cases and none of the validation cases. The 155 development cases were coded blinded to model output and to case-selection origin; they were excluded from the validation sample.

### Independent validation and reference-standard construction

The validation cohort comprised 200 encounters drawn, after the pipeline was frozen, from the 1,514 encounters outside the 155-case development cohort (no overlap, confirmed by accession and patient identifier). Using the same model-consensus enrichment strategy as in development, 50 encounters were selected from the frozen pipeline’s output (never human labels) to over-sample rare findings, prioritizing hemorrhage (including the three remaining model-flagged hemorrhages, all outside the MRI cohort) and basilar stenosis, then CT-head acute findings, MRI acute infarct, and MCA stenosis; 17 of the 50 came from outside the MRI cohort. The other 150 were randomly sampled from the remaining non-development encounters. One hundred encounters (the 50 enriched and 50 random) were independently abstracted by two neurology residents (Reviewer 1 and Reviewer 2), each blinded to the other and to model output; the remaining 100 random encounters were single-read (50 per reviewer). Reviewers were blinded to the origin of each case: random and enriched cases were interleaved in randomized order, and only the principal investigator held the origin key, which was opened after review and adjudication were complete. A single reference standard was built per field: concordant reviewer values were taken as-is, and disagreements were resolved by a blinded vascular neurologist (Reviewer 3). The adjudicator resolved disagreements only and never overrode a concordant cell, so the reference is independent of model output. Accuracy was exact categorical agreement between each model and the adjudicated reference over evaluable cases; not-applicable cells were excluded from numerator and denominator. Because the reference standard is expert abstraction of the same free-text reports, these figures quantify agreement with expert report abstraction (abstraction fidelity) rather than accuracy against imaging ground truth.

### Statistical analysis

Test–retest reproducibility used three same-session runs of Claude Sonnet 4.6 (Fleiss’ κ, exact three-way agreement); temporal reproducibility used a nine-weeks-later GPT-5.4 re-run on the identical cohort (Cohen’s κ, exact agreement). Pairwise inter-model agreement (Cohen’s κ) was computed across the three frontier models, aligned to one encounter order (identifiers 1–1,669). Reproducibility and inter-model agreement are summarized as the mean of field-level κ. For the independent validation, accuracy is the pooled proportion of correctly abstracted Tier 1 cells with 95% Wilson score intervals, and overall agreement is Cohen’s κ with 95% confidence intervals from a case-clustered bootstrap (2,000 resamples); pooled-accuracy Wilson intervals treat cells as independent and do not account for within-encounter clustering, whereas the κ intervals do. Cells with a not-applicable reference or a vessel grade of 9 were excluded; a missing model value against an applicable reference was scored as incorrect. Accuracy is reported for the 150 random encounters (the estimate for the non-enriched remainder), the 50 enriched encounters (rare-finding performance), and the 100 double-read encounters against the adjudicated reference (the comparison benchmarked against inter-reviewer agreement). As a sensitivity analysis, an approximate design-weighted estimate for the 1,514 non-development encounters weighted each random encounter by 1,464/150 and each enriched encounter by 1. Vessel grades were treated as unweighted categorical values across the 0–4 scale; a quadratic-weighted κ sensitivity analysis and per-vessel confusion matrices are provided in the Supplement (Supplementary Tables S2 and S3). Wilson score intervals were also used for prevalence and for sample-size targets.^21^

## Results

### Cohort and imaging utilization

Among 1,669 encounters, CTH was performed in 1,650 (98.9%), CTA head in 1,666 (99.8%), CTA neck in 1,661 (99.5%), and MRI brain in 719 (43.1%); CT perfusion, ascertained from the CTA reports, in 449 (26.9%); and MR angiography, ascertainable only from MRI reports, in 27 of the 719 MRI encounters (3.8%) (Table 1; study overview in Figure 1). The pipeline processed 1,669/1,669 cases with no unrecoverable errors; conditional gating executed a mean of 2.84 of six possible LLM calls per case (the CT-perfusion call in 26.9% of encounters, the MRI calls in 43.1%, and the two infarct-localization calls in 4.6% and 10.7%); each full-cohort run took approximately 4.6 (GPT-5.4), 5.5 (Claude Sonnet 4.6), and 7.7 (Gemini 3.5 Flash) hours on one model backend, roughly 10 to 17 seconds per encounter. Token usage was not logged, so per-case cost is not directly auditable; at public API list prices a full-cohort run is on the order of tens of US dollars per model (an illustrative projection).

### Reproducibility and temporal stability

Across three same-session runs, Tier 1 mean Fleiss’ κ was 0.993 (99.8% exact three-way agreement) and secondary-field κ was 0.991. Re-running the GPT-5.4 configuration on the identical cohort nine weeks later reproduced the output almost exactly (Tier 1 mean κ=0.992, 99.9% exact; secondary κ=0.974), indicating stability across a multi-week interval rather than only within a session; because the models are accessed through a hosted gateway that could in principle be updated between runs, this reflects observed stability of the deployment rather than a guarantee (Table 2, Figure 2).

**Table 2.** Same-session test–retest reproducibility (Claude Sonnet 4.6, three runs), inter-model concordance (three frontier models), and temporal reproducibility (GPT-5.4 re-run nine weeks apart). Unweighted means of field-level statistics. Exact agreement is three-way agreement across the Claude runs (rows 1–2) and run-to-run agreement for the GPT-5.4 re-test (row 3). Vessel cells graded 9 (no CTA report text) are excluded.

| **Field group** | **Test–retest κ** | **Inter-model κ** | **Exact agreement** |
|---|---|---|---|
| Tier 1 (11 fields) | 0.993 | 0.962 | 99.8% |
| Secondary (11 fields) | 0.991 | 0.932 | 99.5% |
| Tier 1 (GPT-5.4 temporal re-test, ~9 wk) | 0.992 | N/A | 99.9% |

**Figure 2.**
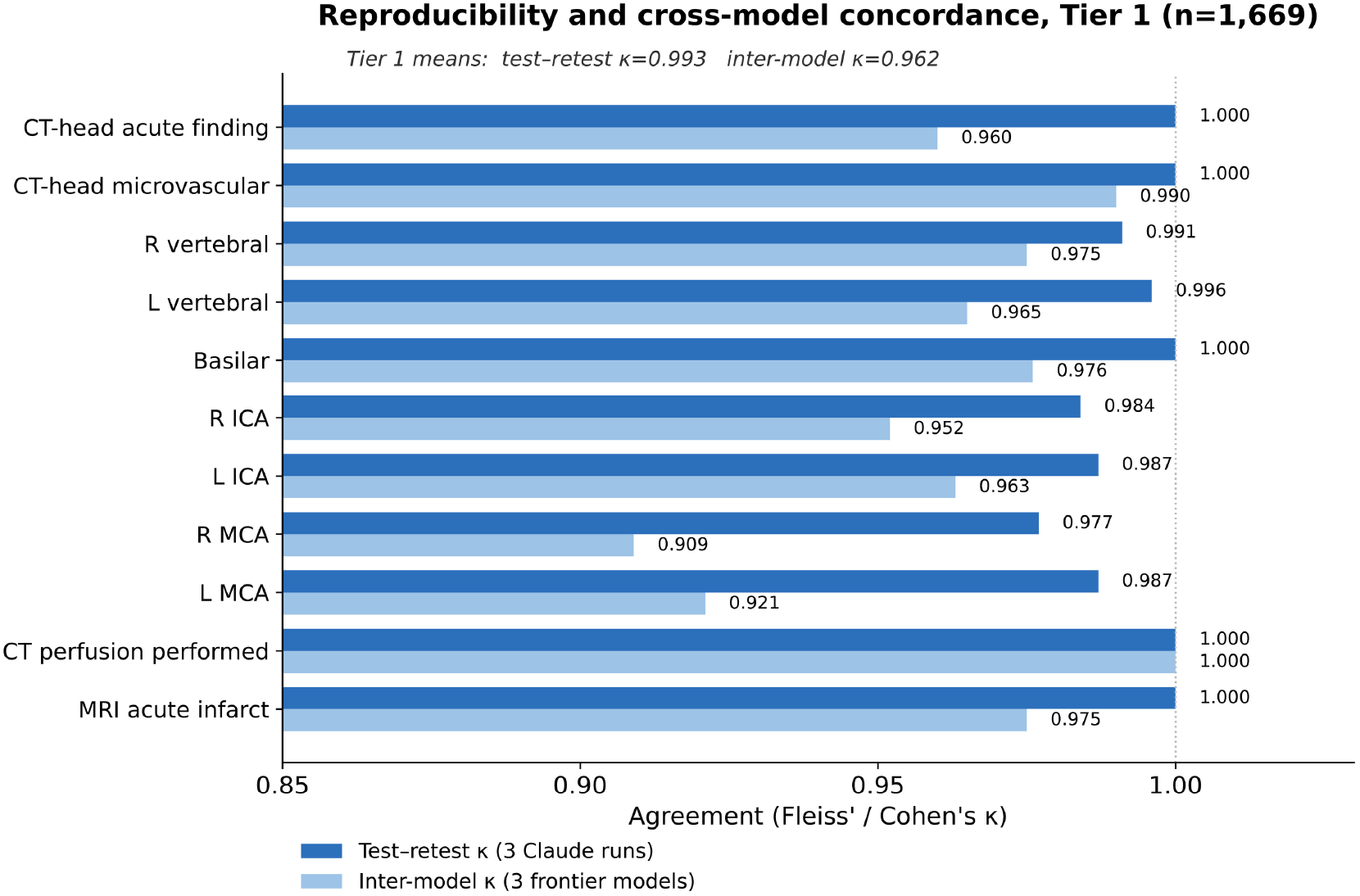
Per-field test–retest reproducibility and inter-model concordance for the eleven Tier 1 fields; agreement is at or near ceiling throughout. The horizontal axis is truncated at 0.85 to resolve near-ceiling differences.

### Inter-model concordance

Run with three independently developed frontier models, mean pairwise agreement was κ=0.962 (Tier 1) and κ=0.932 (secondary), consistent across all three model pairs (Table 2, Figure 2). Because the models were built by different developers, their convergence indicates the structured outputs are reproducible across systems rather than idiosyncratic to one model; because such models may nonetheless share pretraining data and failure modes, this convergence reflects cross-system reproducibility rather than correctness, which is assessed separately against the adjudicated reference.

### Finding prevalence across the full cohort

Tier 1 finding prevalence was highly consistent across the three models, differing by less than one percentage point for every finding (for example, microvascular disease 33.0% [Claude] vs 32.7% [GPT-5.4] vs 32.9% [Gemini]; MRI acute infarct 10.6/10.6/10.7%) (Table 3, Figure 3).

**Table 3.** Prevalence of each Tier 1 finding across the full cohort, by model. n pos is the Claude Sonnet 4.6 count. Denominators are each model’s evaluable encounters: CT head 1,662 (Claude), 1,646 (GPT-5.4), and 1,644 (Gemini); CTA vessels 1,658 for all models (11 encounters had no CTA report text and were coded 9); CT perfusion 1,669; MRI 719, 718, and 709. Claude’s CT-head denominator exceeds the 1,650 structured CT-head studies because, for 17 encounters without a structured CT-head study, it extracted non-contrast head findings from the accompanying CTA report; conversely, 5 encounters with a structured CT-head study were returned as not performed (4 had no report text).

| Tier 1 finding | n pos | Claude % | GPT-5.4 % | Gemini % |
|---|---|---|---|---|
| Acute CT-head finding | 55 | 3.3 | 3.2 | 3.0 |
| Microvascular disease | 549 | 33.0 | 32.7 | 32.9 |
| Right vertebral | 200 | 12.1 | 12.2 | 12.0 |
| Left vertebral | 187 | 11.3 | 11.2 | 11.2 |
| Basilar | 43 | 2.6 | 2.7 | 2.5 |
| Right ICA | 359 | 21.7 | 22.4 | 21.9 |
| Left ICA | 350 | 21.1 | 21.9 | 21.1 |
| Right MCA | 61 | 3.7 | 3.4 | 3.1 |
| Left MCA | 54 | 3.3 | 3.0 | 2.8 |
| CT perfusion performed | 449 | 26.9 | 26.9 | 26.9 |
| MRI acute infarct | 76 | 10.6 | 10.6 | 10.7 |

**Figure 3.**
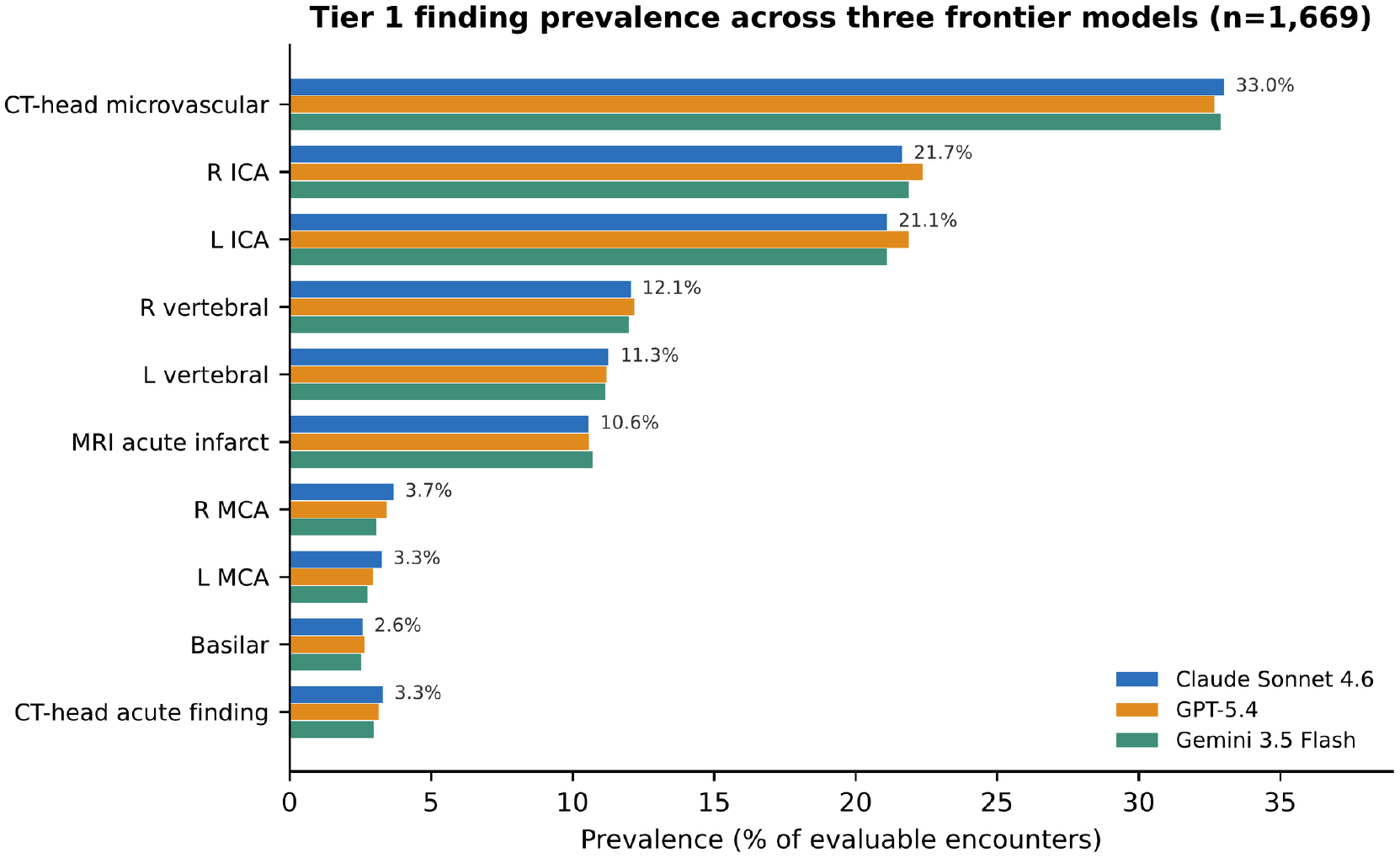
Tier 1 finding prevalence across the three frontier models; estimates differ by <1 percentage point for every finding (the ten clinical Tier 1 findings; CT-perfusion-performed utilization appears in Table 1). This cross-model concordance of prevalence is the population-level counterpart to the field-level agreement in Figure 2 and supports the pipeline output as a stable, model-agnostic prevalence estimate.

### Independent validation against an adjudicated reference

Agreement between the two blinded reviewers was high (pooled Tier 1 Cohen’s κ=0.932, 95% CI 0.909 to 0.954; 95.2% exact across 1,028 Tier 1 cells). Against the adjudicated reference, pooled Tier 1 accuracy on the 100 double-read encounters was 0.982 (95% CI 0.972 to 0.988) for Claude Sonnet 4.6, 0.988 (0.979 to 0.993) for GPT-5.4, and 0.971 (0.959 to 0.980) for Gemini 3.5 Flash. Overall Cohen’s κ was 0.974 (0.961 to 0.986), 0.982 (0.971 to 0.992), and 0.959 (0.936 to 0.979), respectively (bootstrap 95% CIs). On the 100 single-read encounters (coded in equal halves by Reviewer 1 and Reviewer 2, each reviewer’s coding serving as the reference), pooled accuracy was 0.976, 0.975, and 0.975, and across all 200 encounters it was 0.979, 0.982, and 0.973 for the three models (Supplementary Table S1). By sampling stratum, pooled Tier 1 accuracy across the 150 random encounters was 0.983 (0.975 to 0.988) for Claude, 0.983 (0.975 to 0.988) for GPT-5.4, and 0.980 (0.971 to 0.986) for Gemini, and across the 50 enriched encounters 0.968 (0.949 to 0.980), 0.977 (0.961 to 0.987), and 0.955 (0.934 to 0.969) (Table 4; Supplementary Table S1b); the design-weighted estimate was 0.982, 0.983, and 0.979. The 150 random encounters estimate performance among the non-enriched remainder (the design-weighted analysis projects to the full non-development population), whereas the 50 enriched encounters deliberately concentrate the hardest rare-positive cases (reference-positive basilar stenosis in 26 enriched versus 0 random encounters; MRI acute infarct in 17 versus 0) as a stress test for subtle vessel and acute findings. Because the random stratum contains almost no rare positives and the enriched stratum was selected from the models’ own flagged output, these estimates reflect agreement on findings the pipeline reported rather than its sensitivity; this sampling design does not establish population-level sensitivity for individual rare findings. Models assigned grade 9 (no CTA text) to only one double-read encounter (7 vessel-cells) versus 14 and 9 for the two reviewers, so concordance was not inflated by selective exclusion. All three models showed high agreement with the reference (Table 4, Figure 4); inter-reviewer agreement provides context for annotation variability. Per field, Claude reached perfect agreement for microvascular disease, right vertebral, left MCA, CTP status, and MRI acute infarct, and 0.949 to 0.990 for the carotid and basilar vessels; the softest Tier 1 field, the CTH acute finding (0.939 pooled across the 100 double-read encounters; 0.97 in the random and 0.90 in the enriched stratum, Figure 4B), is resolved cross-modally and therefore reflects a harder adjudicated construct. Because the vessel grades are a primary output for reuse, the critical metric is exact field-level agreement with the adjudicated reference. Model-produced vessel grades matched the reference exactly in 99.0% (GPT-5.4; 95% CI 97.9–99.5%), 98.1% (Claude; 96.8– 98.9%), and 96.7% (Gemini; 95.1–97.8%) of vessels; per-vessel exact accuracy ranged 93–100%, was lowest for the internal carotids, and residual errors were almost all a single adjacent grade (within one grade in ≈99% of vessels for each model; Supplementary Table S2 gives model-specific vessel agreement and Table S3 gives Claude’s confusion matrices, including non-adjacent errors). Chance-corrected reliability (unweighted and quadratic-weighted κ) and per-vessel confusion matrices appear in Supplementary Tables S2 and S3. Secondary-field agreement ranged from high (other MRI abnormality 0.960; CTH indeterminate flag 0.969) to low for interpretation-dependent or sparse fields (CTP deficit 0.743; chronic-infarct territory 0.615); all eleven appear in Supplementary Tables S4 and S4b.

**Table 4.** Independent validation, pooled Tier 1 fields, by sampling stratum. Random stratum: all 150 randomly sampled encounters (reference adjudicated for the 50 double-read and a single reviewer for the other 100; 1,527 cells); this is the estimate for the non-enriched remainder. Enriched stratum: 50 encounters selected from pipeline output to over-sample rare findings, all double-read (531 cells). Double-read: the 100 encounters coded by both reviewers against the adjudicated reference; inter-rater agreement is over the 1,028 Tier 1 cells coded by both reviewers, and model comparisons over the 1,045 cells with an applicable reference. Accuracy is the pooled proportion of correctly abstracted Tier 1 cells (Wilson 95% CI); κ is overall Cohen’s κ (case-clustered bootstrap 95% CI). Case-clustered bootstrap intervals for pooled accuracy were of similar width to the Wilson intervals (modestly wider only in the enriched stratum).

| Comparison (Tier 1) | Encounters | Accuracy (95% CI) | κ (95% CI) |
|---|---|---|---|
| Random stratum (150; primary) |  |  |  |
| Claude Sonnet 4.6 | 150 | 0.983 (0.975–0.988) | 0.971 (0.960–0.982) |
| GPT-5.4 | 150 | 0.983 (0.975–0.988) | 0.971 (0.960–0.982) |
| Gemini 3.5 Flash | 150 | 0.980 (0.971–0.986) | 0.966 (0.953–0.978) |
| Enriched stratum (50), adjudicated reference |  |  |  |
| Claude Sonnet 4.6 | 50 | 0.968 (0.949–0.980) | 0.959 (0.939–0.978) |
| GPT-5.4 | 50 | 0.977 (0.961–0.987) | 0.971 (0.953–0.987) |
| Gemini 3.5 Flash | 50 | 0.955 (0.934–0.969) | 0.942 (0.901–0.975) |
| Double-read (100: 50 random + 50 enriched), adjudicated reference |  |  |  |
| Inter-rater (two reviewers) | 100 | 0.952 (exact) | 0.932 (0.909–0.954) |
| Claude Sonnet 4.6 | 100 | 0.982 (0.972–0.988) | 0.974 (0.961–0.986) |
| GPT-5.4 | 100 | 0.988 (0.979–0.993) | 0.982 (0.971–0.992) |
| Gemini 3.5 Flash | 100 | 0.971 (0.959–0.980) | 0.959 (0.936–0.979) |

**Figure 4.**
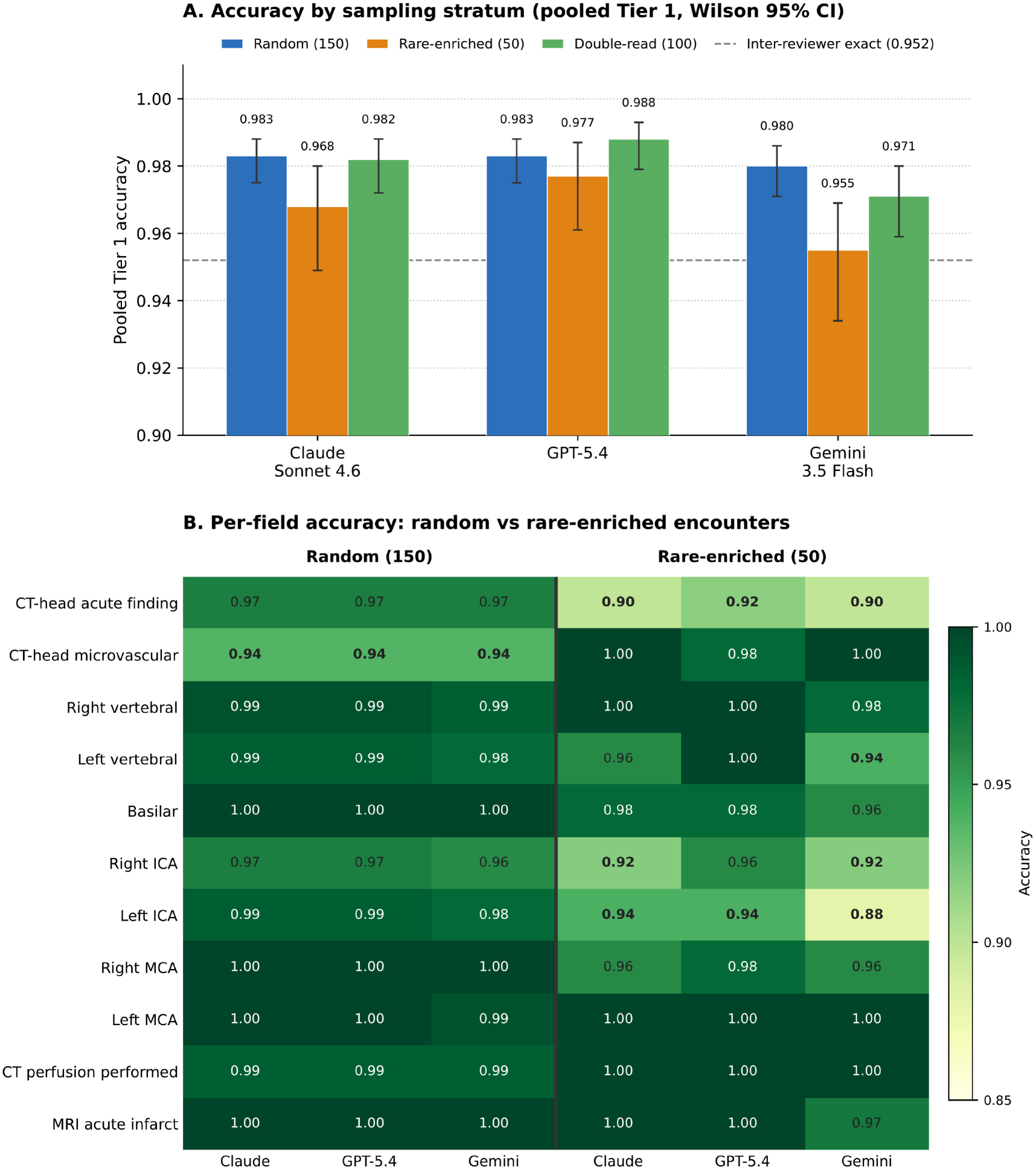
Independent validation by sampling stratum. (A) Pooled Tier 1 accuracy by model for the 150 random encounters (the estimate for the non-enriched remainder), the 50 rare-enriched encounters, and the 100 double-read encounters, each with Wilson 95% confidence intervals; the dashed line marks inter-reviewer exact agreement (0.952); Panel A’s vertical axis is truncated at 0.90 to resolve differences near ceiling. (B) Per-field accuracy for the eleven Tier 1 fields in the random versus the rare-enriched stratum, for all three models. Accuracy is at or near ceiling on the random encounters; the lower values in the enriched stratum fall on the hardest, deliberately over-sampled rare findings (CT-head acute finding and the internal carotid arteries), where positives are concentrated.

Across the program, 355 distinct encounters underwent expert manual review (the 155-case development cohort, which includes the 30 initial prompt-design cases, and the 200 validation encounters). The 155-case development cohort, coded by the separate development reviewer and enriched for rare findings, showed model-versus-human Tier 1 accuracy of 0.974 to 0.975 (κ≈0.94), consistent with the independent-validation result and arguing that agreement reflects schema clarity rather than a model-specific artifact.

## Discussion

A decomposed, modality-specific LLM pipeline abstracted the full four-modality stroke workup across 1,669 ED encounters with very high same-session reproducibility (Tier 1 κ=0.993), stability on a nine-weeks-later re-run (κ=0.992), high cross-developer concordance (κ=0.962), and accuracy against an independent adjudicated reference (0.97–0.99 across three models) at the level of inter-reviewer agreement. Three independent lines of evidence (the same model repeated, three vendors, and models versus humans) converge to within a few percentage points field by field. Model–reference agreement was high and similar in magnitude to the agreement between the two reviewers; because the two comparisons use different reference constructions and evaluable-cell sets, this does not establish human equivalence; residual disagreement likely reflects both report ambiguity and reference-standard noise. These figures quantify the measurement error a downstream analysis would inherit: pooled Tier 1 disagreement with the adjudicated expert abstraction was roughly 1 to 3% in the double-read sample, though error rates varied by field (about 6% for the CT-head acute finding), a modest, now-quantified error budget for future studies to account for. These figures reflect fidelity to expert abstraction of the reports rather than correctness against imaging ground truth; prior work likewise reported high agreement with report abstraction but only moderate agreement with research-grade image scoring.^25^

Reproducibility and robustness under repeated runs are increasingly recognized as under-reported dimensions of clinical LLM extraction^22^; the near-ceiling test–retest, temporal, and cross-model agreement reported here address that gap directly.

We attribute this to three compounding design choices rather than any single innovation: decomposition into narrow, modality-specific LLM calls with fixed categorical schemas and explicit decision rules; iterative, disagreement-driven prompt refinement; and conditional calls with deterministic quality-control overlays. This pattern is consistent with evidence that decomposing a task into narrow sub-tasks reduces cross-field contamination relative to monolithic prompting, though the calls here are fixed and non-interacting rather than autonomous agents. A field-by-field account of successes and struggles (Supplementary Table S5) maps onto the architecture: explicit presence/absence findings, discrete vessel grades, and study ascertainment were captured most reliably, whereas open-vocabulary localization and interpretation-dependent fields (infarct territory, perfusion deficit, carotid grade) remained hardest.^23,24^

A second methodological contribution is the triple-duty use of one modest human-adjudicated cohort: as a development signal that localized ambiguous fields, as a prevalence anchor enriched for rare findings, and as the scarce gold-standard sample in a prevalence-informed enrichment that sized sampling before review. The reviewer coded blinded to model output and to selection origin, so enrichment concentrated expert effort where it was most informative. Because the full cohort was much larger than the development sample, the independent validation could be drawn from genuinely held-out cases; because part of that sample was itself enriched, its accuracy is reported separately for random and enriched encounters.

### Toward population-scale clinical research

These results fit a broader shift in which decomposed LLM systems are moving clinical information extraction from narrow single-task classifiers toward general pipelines that read, reason over, and structure free-text records at scale.^11,16,17,18,19^ The decisive feature for research infrastructure is cost structure: schema design, prompt engineering, and validation are substantial up-front costs (with periodic or change-triggered revalidation), after which the marginal cost per additional case is small, fixed, and dominated by inference tokens rather than expert time. After external multi-site validation, a pipeline of this kind could be applied to cohorts an order of magnitude larger at a low, predictable per-case cost, making previously impractical population-level questions (prevalence, imaging-utilization and practice variation, temporal trends, and the epidemiology of rare but high-consequence findings) tractable, and a cohort feasible to refresh as practice and models evolve. We frame this as a complement to, not a replacement for, expert review: the reproducibility, cross-model, and adjudicated-accuracy reporting shown here, with external validation, are prerequisites for trusting such pipelines as the engine for large research datasets.

### Limitations

This is a single-center, retrospective study; external validation across institutions and reporting conventions remains to be done. The reference standard is expert abstraction of the reports by two neurology residents with vascular-neurology adjudication rather than a neuroradiologist re-read of the source images; agreement therefore reflects fidelity to the documented report, and vessel-grade agreement in particular should be read as agreement with the report rather than with the underlying images. Because MRI was completed in 43.1% of encounters, the CTH acute-finding field is a cross-modally resolved construct in encounters with MRI and a CT-only construct otherwise; on the double-read set, Claude’s accuracy for this field was 97.7% (43/44) without MRI and 90.7% (49/54) with MRI, in small subgroups. CTP and MRA utilization were ascertained from report text rather than structured order fields (CTP from the CTA reports, full cohort; MRA only from MRI reports, 719 MRI encounters). The 50 enriched validation encounters were selected from model-flagged output, so accuracy in that stratum reflects agreement on model-flagged findings and cannot measure how often the model misses a finding. Because the enriched set included every remaining model-flagged basilar stenosis, CT-head acute finding, and hemorrhage, the random stratum contains none of these model-flagged cases; it represents the non-enriched encounters, and the design-weighted estimate approximates the full non-development population. The MRI chronic-infarct location field is free text and not directly comparable across models. The secondary fields that lagged most, chronic-infarct localization and CT-perfusion deficit, are those most exposed to narrative hedging and non-standardized phrasing, where a controlled vocabulary and targeted human review would most help. The study was not designed to validate detection of individual rare acute pathologies. Intracranial hemorrhage in particular was infrequent, and its targeted enrichment drew on the limited pool of model-flagged cases, so pooled accuracy should not be interpreted as validated hemorrhage detection; any hemorrhage-related discordances are retained within the CT-head acute-finding and acute-type fields. Establishing detection performance for rare acute pathologies would require a dedicated, adequately powered study. Finally, the pipeline abstracts report content rather than images and inherits any error in the underlying report.

## Conclusion

A decomposed, modality-specific LLM pipeline produced structured multimodal stroke-workup abstraction that was highly reproducible, temporally stable, portable across the three tested models, and accurate against a rigorously adjudicated reference, at a scale and cost relevant to research data curation. The quantified per-field error rates support research reuse of this dataset with field-specific quality assessment and additional review for weaker or rare fields, and, with external validation, pipelines of this design offer a practical path to large-scale, reproducible clinical research datasets.

## Data Availability

To support reproducibility, the pipeline software (v14.0.3) with institutional API credentials and endpoint removed, the six LLM prompts, and a synthetic test dataset of 75 simulated encounters that exercises the full pipeline are available at Zenodo (https://doi.org/10.5281/zenodo.23824452); the pipeline, the six prompts, and the 75-case synthetic dataset are publicly available (open access) at that link, so reviewers and readers can run the pipeline on the synthetic data.

https://zenodo.org/records/23824452

## Data and code availability

The radiology report text cannot be publicly released: the reports contain identifying information and are governed by the study institutional review board, and full de-identification to a public-release standard was beyond the scope of this work. To support reproducibility, the pipeline software (v14.0.3) with institutional API credentials and endpoint removed, the six LLM prompts, and a synthetic test dataset of 75 simulated encounters that exercises the full pipeline are available at Zenodo (https://doi.org/10.5281/zenodo.23824452); the pipeline, the six prompts, and the 75-case synthetic dataset are publicly available (open access) at that link, so reviewers and readers can run the pipeline on the synthetic data. The de-identified structured model outputs (field values without report text), the adjudicated reference standard, and the analysis code may be shared with qualified investigators on reasonable request, subject to IRB approval and a data-use agreement.

## Use of generative AI

Beyond the LLMs evaluated as the object of study, generative AI (Anthropic Claude) was used under author supervision for pipeline and analysis code and for copy-editing. Study concept and design, all gold-standard coding, interpretation, and scientific content are the authors’ work; all reported values were independently checked against source data. No AI tool met authorship criteria.

