## Supplementary figures and table for "A Reproducible Decomposed Large Language Model Pipeline for Structured Abstraction of Multimodal Neuroimaging Reports: Development and Independent Validation in 1,669 Emergency Department Encounters for Dizziness and Vertigo"

*Contents: Supplementary Tables S1, S1b, S2, S3, S4, S4b, S5. All supplementary content is tabular; there are no supplementary figures. A machine-readable copy is provided as Supplementary_Material.xlsx.*

**Supplementary Table S1.** Pooled Tier 1 accuracy by validation scope (double-read 100, single-read 100, combined 200), by model. Supplementary Table S1b. Pooled Tier 1 accuracy and Cohen’s κ by sampling stratum (random, enriched, double-read, single-read, all 200), by model.

| Scope | Model | Tier1_cells | Pooled_accuracy |
|---|---|---|---|
| double-read (100) | Claude Sonnet 4.6 | 1045 | 0.982 |
| double-read (100) | GPT-5.4 | 1045 | 0.988 |
| double-read (100) | Gemini 3.5 Flash | 1045 | 0.971 |
| single-read (100) | Claude Sonnet 4.6 | 1013 | 0.976 |
| single-read (100) | GPT-5.4 | 1013 | 0.975 |
| single-read (100) | Gemini 3.5 Flash | 1013 | 0.975 |
| combined (200) | Claude Sonnet 4.6 | 2058 | 0.979 |
| combined (200) | GPT-5.4 | 2058 | 0.982 |
| combined (200) | Gemini 3.5 Flash | 2058 | 0.973 |

**Supplementary Table S1b.** Accuracy and κ each shown as point estimate (95% CI).

| Stratum | Model | Enc. | Cells | Accuracy (95% CI) | Cohen's κ (95% CI) |
|---|---|---|---|---|---|
| Double-read (100) | Claude | 100 | 1045 | 0.982 (0.972–0.988) | 0.974 (0.961–0.986) |
| Double-read (100) | GPT-5.4 | 100 | 1045 | 0.988 (0.979–0.993) | 0.982 (0.971–0.992) |
| Double-read (100) | Gemini | 100 | 1045 | 0.971 (0.959–0.98) | 0.959 (0.936–0.979) |
| DR, random (50) | Claude | 50 | 514 | 0.996 (0.986–0.999) | 0.994 (0.984–1) |
| DR, random (50) | GPT-5.4 | 50 | 514 | 0.998 (0.989–1) | 0.997 (0.99–1) |
| DR, random (50) | Gemini | 50 | 514 | 0.988 (0.975–0.995) | 0.981 (0.962–0.997) |
| DR, enriched (50) | Claude | 50 | 531 | 0.968 (0.949–0.98) | 0.959 (0.939–0.978) |
| DR, enriched (50) | GPT-5.4 | 50 | 531 | 0.977 (0.961–0.987) | 0.971 (0.953–0.987) |
| DR, enriched (50) | Gemini | 50 | 531 | 0.955 (0.934–0.969) | 0.942 (0.901–0.975) |
| Single-read (100) | Claude | 100 | 1013 | 0.976 (0.965–0.984) | 0.959 (0.943–0.975) |
| Single-read (100) | GPT-5.4 | 100 | 1013 | 0.975 (0.964–0.983) | 0.958 (0.941–0.974) |
| Single-read (100) | Gemini | 100 | 1013 | 0.975 (0.964–0.983) | 0.958 (0.941–0.974) |
| All random (150) | Claude | 150 | 1527 | 0.983 (0.975–0.988) | 0.971 (0.96–0.982) |
| All random (150) | GPT-5.4 | 150 | 1527 | 0.983 (0.975–0.988) | 0.971 (0.96–0.982) |
| All random (150) | Gemini | 150 | 1527 | 0.98 (0.971–0.986) | 0.966 (0.953–0.978) |
| All (200) | Claude | 200 | 2058 | 0.979 (0.972–0.984) | 0.968 (0.958–0.977) |
| All (200) | GPT-5.4 | 200 | 2058 | 0.982 (0.975–0.987) | 0.972 (0.962–0.981) |
| All (200) | Gemini | 200 | 2058 | 0.973 (0.965–0.979) | 0.959 (0.944–0.972) |

*DR = double-read. Design-weighted accuracy for the 1,514 non-development encounters: Claude 0.982, GPT-5.4 0.983, Gemini 0.979.*

**Supplementary Table S2.** Per-vessel agreement with the adjudicated reference (100 double-read): exact accuracy (95% Wilson CI), misclassification rate, within-one-grade agreement, unweighted and quadratic-weighted Cohen's kappa (bootstrap CI), and present-versus-absent kappa.

| Model | Vessel | N | Exact acc. % (95% CI) | Within-1 % | Unwtd κ | Quad-wtd κ (95% CI) | P/A κ |
|---|---|---|---|---|---|---|---|
| Claude Sonnet 4.6 | Right vertebral | 99 | 100 (96.3-100.0) | 100 | 1 | 1 (1.000-1.000) | 1 |

| | | | | | | | |
|---|---|---|---|---|---|---|---|
| Claude Sonnet 4.6 | Left vertebral | 99 | 98 (92.9-99.4) | 99 | 0.952 | 0.966 (0.891-1.000) | 0.973 |
| Claude Sonnet 4.6 | Basilar | 99 | 99 (94.5-99.8) | 100 | 0.977 | 0.994 (0.978-1.000) | 0.974 |
| Claude Sonnet 4.6 | Right ICA | 99 | 94.9 (88.7-97.8) | 98 | 0.904 | 0.863 (0.662-0.990) | 0.932 |
| Claude Sonnet 4.6 | Left ICA | 99 | 97 (91.5-99.0) | 100 | 0.942 | 0.987 (0.972-1.000) | 1 |
| Claude Sonnet 4.6 | Right MCA | 99 | 98 (92.9-99.4) | 100 | 0.868 | 0.973 (0.916-1.000) | 1 |
| Claude Sonnet 4.6 | Left MCA | 99 | 100 (96.3-100.0) | 100 | 1 | 1 (1.000-1.000) | 1 |
| GPT-5.4 | Right vertebral | 99 | 100 (96.3-100.0) | 100 | 1 | 1 (1.000-1.000) | 1 |
| GPT-5.4 | Left vertebral | 99 | 100 (96.3-100.0) | 100 | 1 | 1 (1.000-1.000) | 1 |
| GPT-5.4 | Basilar | 99 | 99 (94.5-99.8) | 100 | 0.977 | 0.994 (0.979-1.000) | 0.974 |
| GPT-5.4 | Right ICA | 99 | 98 (92.9-99.4) | 100 | 0.962 | 0.99 (0.975-1.000) | 1 |
| GPT-5.4 | Left ICA | 99 | 97 (91.5-99.0) | 100 | 0.942 | 0.987 (0.973-1.000) | 1 |
| GPT-5.4 | Right MCA | 99 | 99 (94.5-99.8) | 100 | 0.934 | 0.987 (0.947-1.000) | 1 |
| GPT-5.4 | Left MCA | 99 | 100 (96.3-100.0) | 100 | 1 | 1 (1.000-1.000) | 1 |
| Gemini 3.5 Flash | Right vertebral | 99 | 98 (92.9-99.4) | 100 | 0.957 | 0.993 (0.981-1.000) | 0.975 |
| Gemini 3.5 Flash | Left vertebral | 99 | 96 (90.1-98.4) | 99 | 0.902 | 0.959 (0.886-0.997) | 0.945 |
| Gemini 3.5 Flash | Basilar | 99 | 98 (92.9-99.4) | 100 | 0.953 | 0.989 (0.965-1.000) | 0.948 |
| Gemini 3.5 Flash | Right ICA | 99 | 94.9 (88.7-97.8) | 98 | 0.901 | 0.916 (0.791-0.990) | 0.931 |
| Gemini 3.5 Flash | Left ICA | 99 | 92.9 (86.1-96.5) | 96 | 0.858 | 0.916 (0.815-0.978) | 0.906 |
| Gemini 3.5 Flash | Right MCA | 99 | 98 (92.9-99.4) | 98 | 0.85 | 0.781 (0.320-1.000) | 0.847 |
| Gemini 3.5 Flash | Left MCA | 99 | 99 (94.5-99.8) | 100 | 0.92 | 0.985 (0.852-1.000) | 0.918 |

**Supplementary Table S3.** Per-vessel 5x5 confusion matrices (reference x model grade, 0-4), Claude Sonnet 4.6, 100 double-read.

**Right vertebral (rows = reference grade; columns = model grade, 0–4)**

| ref\model | 0 | 1 | 2 | 3 | 4 |
|---|---|---|---|---|---|
| 0 | 70 | 0 | 0 | 0 | 0 |
| 1 | 0 | 10 | 0 | 0 | 0 |
| 2 | 0 | 0 | 5 | 0 | 0 |
| 3 | 0 | 0 | 0 | 8 | 0 |
| 4 | 0 | 0 | 0 | 0 | 6 |

**Left vertebral (rows = reference grade; columns = model grade, 0–4)**

| ref\model | 0 | 1 | 2 | 3 | 4 |
|---|---|---|---|---|---|
| 0 | 74 | 1 | 0 | 0 | 0 |
| 1 | 0 | 7 | 0 | 0 | 1 |
| 2 | 0 | 0 | 4 | 0 | 0 |
| 3 | 0 | 0 | 0 | 6 | 0 |
| 4 | 0 | 0 | 0 | 0 | 6 |

**Basilar (rows = reference grade; columns = model grade, 0–4)**

| ref\model | 0 | 1 | 2 | 3 | 4 |
|---|---|---|---|---|---|
| 0 | 72 | 1 | 0 | 0 | 0 |
| 1 | 0 | 9 | 0 | 0 | 0 |
| 2 | 0 | 0 | 11 | 0 | 0 |
| 3 | 0 | 0 | 0 | 5 | 0 |
| 4 | 0 | 0 | 0 | 0 | 1 |

**Right ICA (rows = reference grade; columns = model grade, 0–4)**

| ref\model | 0 | 1 | 2 | 3 | 4 |
|---|---|---|---|---|---|
| 0 | 64 | 0 | 0 | 0 | 1 |
| 1 | 1 | 12 | 0 | 0 | 0 |
| 2 | 0 | 1 | 12 | 0 | 0 |
| 3 | 1 | 0 | 0 | 6 | 1 |
| 4 | 0 | 0 | 0 | 0 | 0 |

**Left ICA (rows = reference grade; columns = model grade, 0–4)**

| ref\model | 0 | 1 | 2 | 3 | 4 |
|---|---|---|---|---|---|
| 0 | 66 | 0 | 0 | 0 | 0 |
| 1 | 0 | 12 | 0 | 0 | 0 |
| 2 | 0 | 1 | 10 | 0 | 0 |
| 3 | 0 | 0 | 0 | 7 | 1 |
| 4 | 0 | 0 | 0 | 1 | 1 |

**Right MCA (rows = reference grade; columns = model grade, 0–4)**

| ref\model | 0 | 1 | 2 | 3 | 4 |
|---|---|---|---|---|---|
| 0 | 91 | 0 | 0 | 0 | 0 |
| 1 | 0 | 3 | 0 | 0 | 0 |
| 2 | 0 | 1 | 1 | 0 | 0 |
| 3 | 0 | 0 | 0 | 2 | 1 |
| 4 | 0 | 0 | 0 | 0 | 0 |

**Left MCA (rows = reference grade; columns = model grade, 0–4)**

| ref\model | 0 | 1 | 2 | 3 | 4 |
|---|---|---|---|---|---|
| 0 | 92 | 0 | 0 | 0 | 0 |
| 1 | 0 | 3 | 0 | 0 | 0 |
| 2 | 0 | 0 | 2 | 0 | 0 |
| 3 | 0 | 0 | 0 | 1 | 0 |
| 4 | 0 | 0 | 0 | 0 | 1 |

**Supplementary Table S4.** All eleven secondary (Tier 2) fields: N, accuracy, and kappa against the adjudicated reference (Claude Sonnet 4.6, 100 double-read). MRA inclusion is scored only among encounters with MRI.

| Secondary (Tier 2) field | N | Accuracy | Cohen's κ |
|---|---|---|---|
| MRI chronic infarct | 54 | 0.963 | 0.914 |
| MRI acute territory | 14 | 0.929 | 0.884 |
| MRI other abnormality | 50 | 0.96 | 0.728 |
| CT-head acute indeterminate | 97 | 0.969 | 0.784 |
| MRI acute bilateral | 18 | 0.889 | 0.76 |
| MRI other abnormality type | 40 | 0.925 | 0.677 |
| MRI chronic bilateral | 20 | 0.75 | 0.587 |
| CT-head acute type | 19 | 0.737 | 0.615 |
| CT perfusion deficit | 35 | 0.743 | 0.512 |
| MRA included | 51 | 0.961 | 0.485 |
| MRI chronic territory | 13 | 0.615 | 0.467 |

*Note: fields are scored only among double-read encounters with an evaluable reference for that field. Of the 54 double-read encounters with MRI, MRA was assessable in 51 (1 reference-positive), since MRA can be ascertained only from MRI reports.*

**Supplementary Table S4b.** Description of each secondary (Tier 2) field.

| Tier 2 field | Description |
|---|---|
| CTH_Acute_Type | Type of acute CT-head finding (ischemic infarct, hemorrhage, or mass/edema) |
| CTH_Acute_Indeterminate | Flag for an indeterminate or equivocal acute CT-head finding |
| CTP_Deficit | Presence of a perfusion deficit on CT perfusion |
| MRI_Chronic_Infarct | Chronic infarct present on MRI |
| MRI_Acute_Territory | Vascular territory of the acute infarct |
| MRI_Acute_Bilateral | Bilateral acute infarct |

| | |
|---|---|
| MRI_Chronic_Territory | Vascular territory of the chronic infarct |
| MRI_Chronic_Bilateral | Bilateral chronic infarct |
| MRI_Other_Abnormality | Other (incidental or non-stroke) MRI abnormality present |
| MRI_Other_Abnormality_Type | Category of the other MRI abnormality |
| MRI_MRA_Included | MR angiography acquired with the MRI brain study |

**Supplementary Table S5.** Field-by-field account of pipeline strengths and failure modes, with the corresponding role of the decomposed modular and deterministic design.

| Category | Representative fields | Validation performance | Pattern / failure mode | How the decomposed modular + deterministic design addressed it |
|---|---|---|---|---|
| STRENGTH: explicit presence/absence | Microvascular disease; MRI acute infarct; CT-perfusion performed; MRA included | Human-reference accuracy 96-100%; full-cohort CTP/MRA consistency checks reported separately | These are stated explicitly in the report as present or absent | Modality-specific LLM calls with fixed yes/no schemas plus a deterministic negation/QC layer captured explicit statements and suppressed unnegated-language errors |
| STRENGTH: discrete vessel grading | Vertebral, basilar, and middle cerebral artery stenosis | Exact grade accuracy 98-100% (lead models) | Ordinal 0-4 grade stated per named vessel | A dedicated CTA vessel-grading call with a fixed 0-4 scale per vessel reduced cross-field contamination and standardized the ordinal mapping |
| STRENGTH: cross-modal reconciliation | CT-head acute finding | CT-head acute-finding accuracy 0.939 (double-read) | An indeterminate initial CT is later clarified by MRI | The deterministic confidence/QC layer reconciled an indeterminate CT to acute when MRI confirmed infarction, rather than forcing a single-pass CT read |
| FAILURE MODE: multi-level vessels | Internal carotid artery grade | ~3-5% field error; the most frequent vessel disagreement | Carotid disease described at multiple segments (bulb, cavernous, petrous) and as tandem lesions, hard to collapse to one grade | Vessel-specific prompting reduced but did not eliminate ambiguity; a per-segment carotid schema is a planned refinement |
| FAILURE MODE: open-vocabulary localization | Chronic- and acute-infarct territory; other-abnormality type | Highest inter-model disagreement (2.5-4.4%) and error (up to ~38% on small n) | Non-standardized anatomical phrasing and an open label space | Pre-specified as secondary/exploratory; a controlled vocabulary for territory and abnormality type is planned |
| FAILURE MODE: interpretation-dependent | CT-perfusion deficit | ~26% field error | Deficits hedged in the report (subtle, equivocal, artifact vs deficit) | Flagged as exploratory; benefits most from targeted human review |
| FAILURE MODE: sparse / indeterminate | CT-head acute type (ischemic vs hemorrhage vs mass) | ~26% error on a small, sparse field | Initial CT frequently indeterminate for the specific type | An acute-indeterminate flag plus cross-modal reconciliation (MRI) partially resolved the type where MRI was available |
| FAILURE MODE: diagnostic boundary | Chronic infarct vs microvascular disease vs chronic hemorrhage | Chronic-infarct accuracy 0.963 (κ 0.914) | Overlapping radiologic descriptors at a recognized inter-observer boundary | Resident disagreements at this boundary were resolved by blinded vascular-neurology adjudication (blind to model output, based on resident codings only), consistent with the independent reference; residual difficulty reflects intrinsic inter-observer ambiguity rather than a pipeline defect |
